# Internet and mental health during the COVID-19 pandemic: Evidence from the UK

**DOI:** 10.1101/2022.04.01.22273299

**Authors:** Climent Quintana-Domeque, Jingya Zeng, Xiaohui Zhang

## Abstract

With the COVID-19 pandemic, the Internet has become a key player in the daily lives of most people. We investigate the relationship between mental health and internet use frequency and purpose six months after the first lockdown in the UK, September 2020. Using data from the UK Household Longitudinal Study on the 12-item General Health Questionnaire and the Internet use module, and controlling for sociodemographic characteristics and personality traits, we find that older individuals (aged 59 or above) have a lower internet use frequency (twice a day or less). Younger women use the Internet for social purposes more than men do, while younger men use the Internet for leisure-and-learning purposes more than women and older men do. Both high frequency internet use and use for social purposes appear to be a protective factor for social dysfunction. Interestingly, high internet use is a protective factor for social dysfunction among younger women, but a risk factor for psychological distress among younger men. Finally, while leisure-and-learning purpose is a protective factor for social dysfunction among younger women, it is a risk factor for social dysfunction among younger men.

## 1 Introduction

The COVID-19 pandemic and the measures to contain the transmission of the virus have transformed our daily routines. For many people, the Internet has become a central part of their lives, from online education to working-from-home. At the same time, the mental wellbeing of the population has been negatively affected by the pandemic (Proto & Quintana-Domeque, 2021; Quintana-Domeque & Proto, 2022), and this deterioration has been heterogeneous among demographic groups, with younger adults and women being disproportionately affected (Banks & Xu, 2020). Our focus in this paper is on the link between the Internet and mental health during the COVID-19 pandemic, with a particular focus on whether such a relationship varies between younger and older adults, and between men and women.

We use data from the UK Household Longitudinal Study to answer three questions: First, do age and gender predict internet use frequency and purpose? Second, do internet use frequency and purpose predict mental health? Third, do internet use frequency and purpose predict mental health differently by age and gender? In order to answer these questions we use three different measures of mental health (Likert GHQ-12 score (0-36), psychological distress score, and social dysfunction score), a measure of internet use frequency (low –twice a day or less– vs. high – more than twice a day), three scores of internet use purpose (functional, social, and leisure-and-learning), and a host of control variables, including personality traits, whose relevance in understanding the consequences of the COVID-19 pandemic on mental health has been recently shown (Proto & Zhang, 2021).

Our multiple regression analysis reveals the following main findings: first, older individuals use the Internet less frequently; second, younger women use the Internet for social purposes more than men do, while younger men use the Internet for leisure-and-learning purposes more than women and older men do; third, high frequency internet use appears to be a protective factor for social dysfunction; fourth, the use of internet for social purposes appears to be a protective factor for social dysfunction; fifth, internet use is a protective factor for social dysfunction among younger women, but a risk factor for psychological distress among younger men; finally, while using the Internet for leisure-and-learning purposes is a protective factor for social dysfunction among younger women, it is a risk factor for social dysfunction among younger men.

The relationship between internet and mental health has been studied previously in economics and psychology. Golin (2021), who provides a detailed summary of the literature on the relationship between internet and mental health, investigates the causal effect of broadband internet access on the mental health of adults aged 17-59 in Germany. Inspired by the work of Falck et al. (2014), her identification strategy to deal with both unobservable determinants of mental health and behaviours and reverse causality is based on a natural experiment which exploits technological features of the German telecommunication network. Her findings suggest that broadband internet has negative effects on women’s mental health but not on men’s. To the best of our knowledge, Golin (2021) is the first study to provide convincing evidence of a causal relationship of internet (broadband access) on validated measures of mental health amongst adults.

We depart from Golin (2021) in four different ways. First, the populations under study are different. Our focus is the UK, not Germany, and our sample is restricted to individuals aged 23-93, not 17-59. Second, our analysis focuses on the COVID-19 pandemic period, a period when the Internet has become a key player in the daily lives of most people. Third, as Golin (2021) acknowledges, the effect of the Internet use on mental health depends on the type of activities that are carried out online, but her paper cannot speak about that. Our study relies on rich Internet data on both use frequency and purposes in September 2020, after contributing suggested content that became the Internet use module of the September 2020 COVID-19 wave study (pp. 123-127, https://www.understandingsociety.ac.uk/sites/default/files/downloads/documentation/covid-19/questionnaires/wave-5/W5-covid-19-questionnaire.pdf). Finally, our cross-sectional study is mainly descriptive, while Golin (2021) uses time variation and focuses on causality by exploiting technological features of the German telecommunication network. Nevertheless, we investigate the plausibility of our analysis capturing the causal effect of internet use on mental health by using Oster (2019)’s bounds analysis.

The next section describes the data sources and the variables used in the analysis, including those obtained after using principal component analysis (functional, leisure-and-learning, and social purposes scores of internet use) and exploratory factor analysis (psychological distress and social dysfunction scores of mental health dimensions). Section 3 contains the results of our multiple regression analysis. In Section 4 we account for multiple testing and use a bounds approach to assess whether our estimated conditional associations (between internet frequency and mental health) can be given a causal interpretation. Finally, section 5 concludes.

## 2 Data sources, variables and descriptive statistics

Our main data source is the UK Household Longitudinal Study (UKHLS).^1^ The UKHLS, also known as Understanding Society, is a large national probability-based household panel survey, involving over 100,000 individuals in 40,000 households in the United Kingdom since wave 1 (2009-2011) (Institute for Social and Economic Research, 2020). The UKHLS provides high-quality longitudinal data on subjects such as social life, education, employment, personality, health and wellbeing. All members of households aged 16+ who participated in waves 8 or 9 of the UKHLS main survey were invited to participate in the COVID-19 study. The COVID-19 study is an integral part of the UKHLS, which is a panel study on experiences and how the UK population reacts to the COVID-19 pandemic (Institute for Social and Economic Research, 2021). The first wave of the COVID-19 survey was in April 2020, and the last one in March 2021.

Analyses are conducted using Stata (version 17). All analyses account for survey design and sample weights using the svyset command in Stata, so that we adjust for unequal selection probabilities and differential non-response to ensure the results are representative of the UK population.^2^

The key variables in our analysis can be classified into three groups: internet use (frequency and purpose), mental health, and sociodemographic and personality characteristics.

### 2.1 Internet use frequency and internet use purpose

All waves of the COVID-19 study collect information on household access to the Internet from home. Moreover, we contributed suggested content that became the Internet use module of the September 2020 COVID-19 wave study. The special Internet Use Module collects detailed internet use information, including frequency of using the Internet and frequency of ten different online activities (browsing websites, email, looking at social media, posting on social media, online buying, online banking, gaming, streaming videos, streaming music, and education). For this reason, the September 2020 COVID-19 wave study is our main data resource for internet usage. We use two types of internet measures: internet use frequency and internet use purpose.

#### Internet use frequency

Respondents were asked how often they use the Internet for their personal use, frequency from “Almost all of the time” to “Never use” (https://www.understandingsociety.ac.uk/documentation/covid-19/dataset-documentation/variable/netpusenew). The frequency of using the Internet is categorised into a binary variable to represent low (twice a day or less) and high levels (more than twice a day) of internet use.

#### Internet use purpose

Respondents were asked how often they use the Internet for personal use to do a specific online activity. There are ten online activities in the questionnaire, including browsing websites, email, looking at social media, posting on social media, online buying, online banking, gaming, streaming videos, streaming music and education (pp. 123-127, https://www.understandingsociety.ac.uk/sites/default/files/downloads/documentation/covid-19/questionnaires/wave-5/W5-covid-19-questionnaire.pdf). Each activity is re-coded from 6 (every day) to 1 (never), where higher scores indicate personal internet use for the activity at the frequency level. Several online activities are highly correlated with each other. For instance, the correlations between browsing websites and email, looking at social media and posting on social media, and streaming videos and streaming music are 0.735, 0.603 and 0.698, respectively. We use principal component analysis (PCA) with Promax rotation to extract the important information from ten online activities and reduce the dimensionality of the data set (Abdi & Williams, 2010; Bro & Smilde, 2014). Respondents in the September survey with valid answers on ten online activities are used for PCA.

The PCA indicated a three-component solution, explaining 64% of the total variance in online activities (component 1 eigenvalue = 4.18, component 2 eigenvalue = 1.20, component 3 eigenvalue = 1.03). Variables loading heavily on the first component (26.6% of the variance) are browsing websites, email, online banking and online buying. Variables loading heavily on the second component (20.8% of the variance) are streaming videos, streaming music and online education. Variables loading heavily on the third component (17.4% of the variance) are looking at social media, posting on social media and gaming.

We label the components “Functional purpose”, “Leisure-and-learning purpose” and “Social purpose”, respectively and use them as a measurement of respondents’ internet use purposes (see PCA and the results in Appendix A). Table 1 provides the descriptive statistics of the Internet purposes scores. Higher scores indicate higher internet use frequency for that purpose. Scores are standardised in the final sample.

**Table 1:** Descriptive statistics of internet use purposes scores

| Variable | N | Mean | Std.Dev. | Min | Max |
| --- | --- | --- | --- | --- | --- |
| Functional purpose score | 12,811 | 6.12e-09 | 1.60 | -5.64 | 2.45 |
| Leisure-and-learning purpose score | 12,811 | 1.76e-10 | 1.45 | -2.19 | 3.42 |
| Social purpose score | 12,811 | 1.60e-09 | 1.30 | -2.73 | 2.37 |

Note: This table shows descriptive statistics of internet use purposes of 12,811 respondents with valid answers on ten online activity questions in the September survey.

### 2.2 Mental health metrics

The 12-item General Health Questionnaire (GHQ-12) is included in every wave of the UKHLS main survey and COVID-19 survey. The GHQ-12 is a reliable and valid self-administered questionnaire designed to identify minor psychiatric disorders in community samples (Goldberg, 1988), a widely accepted indicator of mental wellbeing (Goldberg et al., 1997), and has been used to assess the impact of the COVID-19 pandemic on mental health in the UK (Banks & Xu, 2020; Proto & Quintana-Domeque, 2021; Proto & Zhang, 2021; Quintana-Domeque & Proto, 2022). The twelve items collect information about how individuals feel about themselves on concentration, anxiety-based insomnia, capability in coping, ability to enjoy day-to-day activities, confidence, being under strain, feeling depressed and unhappiness, among others over the last few weeks on four-point Likert scale (https://doi.org/10.1371/journal.pone.0244419.s001).

We use three measures based on the GHQ-12. The first measure is the Likert GHQ-12, a continuous Likert scale that sums the 12 items of the GHQ. Each item scores from 0 (better than usual) to 3 (much worse than usual), and the Likert GHQ-12 score ranges from 0 (best mental wellbeing) to 36 (worst mental wellbeing).

The other two measures exploit the potential multidimensional properties of the GHQ-12 score (Graetz, 1991; Romppel et al., 2013; Griffith & Jones, 2019). We follow the literature using exploratory factor analysis (EFA) with Varimax rotation (Williams et al., 2010) to identify dimensions of the GHQ-12 in the September survey. Varimax rotation produces factor structures that are uncorrelated and simplifies the interpretation of the factors by minimizing the number of variables that have high loadings on each factor (Williams et al., 2010). 12,419 respondents in the September survey with valid answers on the GHQ-12 questionnaire are used for EFA. The EFA yielded a two-factor solution, explaining 73.73% of the total variance in items (factor 1 eigenvalue = 7.67 and factor 2 eigenvalue = 1.17). We label the factors *psychological distress* and *social dysfunction* respectively (see factor analysis and the results in Appendix B).

The *psychological distress* score is related to anxiety and depression, more precisely, anxiety-based insomnia (item 2 of the GHQ-12), under strain (item 5), problem overcoming difficulties (item 6), depression (item 9), lose confidence (item 10) and believe worthless (item 11). The *social dysfunction* score is related to the ability to perform daily activities and to cope with everyday problems, more precisely, concentration (item 1 of the GHQ-12), playing a useful role (item 3), decision making (item 4), ability to enjoy day-to-day activities (item 7), face up to problems (item 8), and unhappiness (item 12). Table 2 provides the descriptive statistics of the psychological distress and social dysfunction scores. Higher scores indicate higher level of psychological distress or social dysfunction. Scores are standardised in the final sample.

**Table 2:** Descriptive statistics of psychological distress and social dysfunction scores

|  | N | Mean | Std.Dev. | Min | Max |
| --- | --- | --- | --- | --- | --- |
| Psychological distress score | 12,419 | -.001 | 0.963 | -2.731 | 3.897 |
| Social dysfunction score | 12,419 | -.003 | 0.942 | -3.998 | 4.828 |

Note: This table shows descriptive statistics of extracted two factors of the GHQ-12 of 12,419 respondents with valid answers on GHQ-12 questionnaire in the September survey.

### 2.3 Control variables

We use the following set of control variables: age, gender (=1 if female, =0 if male), ethnic minority (=1 if Black, Asian or other ethnic minority, = 0 if White British), education (measured in 2017-2019), marital status, household size, employment status, household income, health status, COVID-19 symptoms, risky behaviours (smoking and drinking), physical exercise, geographical location, pre-pandemic mental wellbeing (measured in 2017-2019), mental wellbeing in July 2020, and personality traits (measured in 2011-2013). Table C1 in Appendix C contains the definition of each variable.

In order to use all this information, we match wave 3 (2011-13), wave 9 (2017-19), the July 2020 wave and the September 2020 wave, and end up with a sample of 6,589 individuals aged 23-93 (5,980 of them have non-zero cross-sectional sample weights in the September 2020 wave). The original sample size with information on the GHQ-12 is 10,267 (and with information on personality traits is 8,589-8,590), while the final sample size with information on all the relevant (including control) variables is 5,980. Thus the final sample represents 58% (70%) of the original sample. Table D1 in Appendix D reveals no differences between the original and the final samples in terms of internet use frequency, the fraction of men and women, the fraction of individuals who have/had COVID-19 symptoms, the percentage of individuals who drink heavily, the fraction of individuals who do regular exercise, and the big 5 personality traits. However, we find statistically differences at the 5% or lower significance level in terms of mental health, age, ethnicity, education, marital status, employment, income, and smoking status.^3^

### 2.4 Descriptive statistics

Table E1 in Appendix E provides a description of the average characteristics of the individuals in our final sample. The average age is 54.65 (SD=14.66), 53.8% of them are women, 91.6% are White British, 60.0% of them are employed, and 47.2% have a higher education degree. In terms of mental health and internet use, the average GHQ-12 score is 11.85 (5.56), and 74% of individuals use the Internet more than twice a day.

In Figure 1 we plot the average mental health metrics by internet use frequency. Individuals who report a high internet use frequency (more than twice a day) tend to report a larger GHQ-12 score (11.96, 95% CI: 11.67, 12.24) than individuals reporting a low internet use frequency (11.54, 95% CI: 11.05, 12.04). While this difference is not significant, individuals reporting a low internet use frequency tend to significantly exhibit a lower psychological distress score but perform worse in terms of the social dysfunction score.

**Figure 1:**
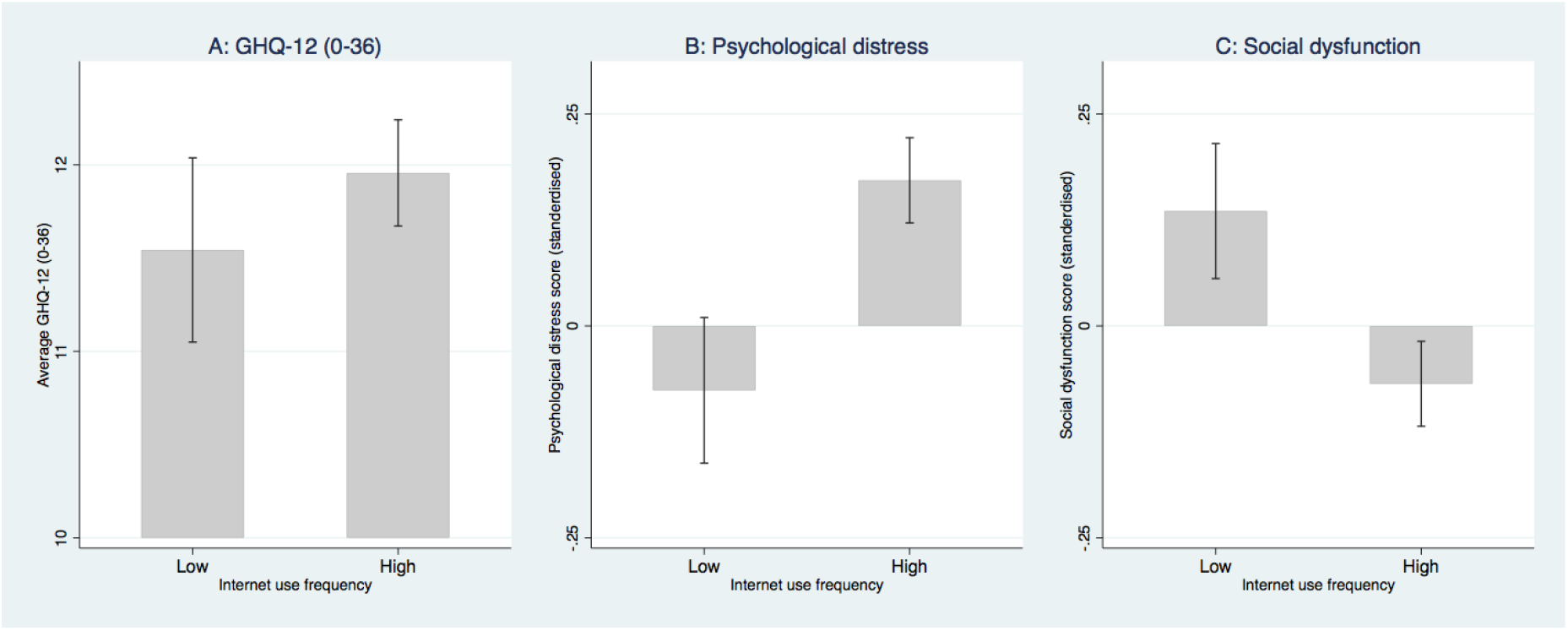
Mental health metrics by internet use frequency

Figure 2 plots average mental health metrics by different levels of the functional purpose score (low (< 50%) –below the median score– vs. high (≥ 50%) – above the median score). There is a positive gradient between the average psychological distress score and functional purpose score, and a negative gradient between the average social dysfunction score and functional purpose score. Similar qualitative gradients are observed when plotting the average psychological distress and social dysfunction scores against leisure-and-learning (Figure 3) and social (Figure 4) purposes scores.

**Figure 2:**
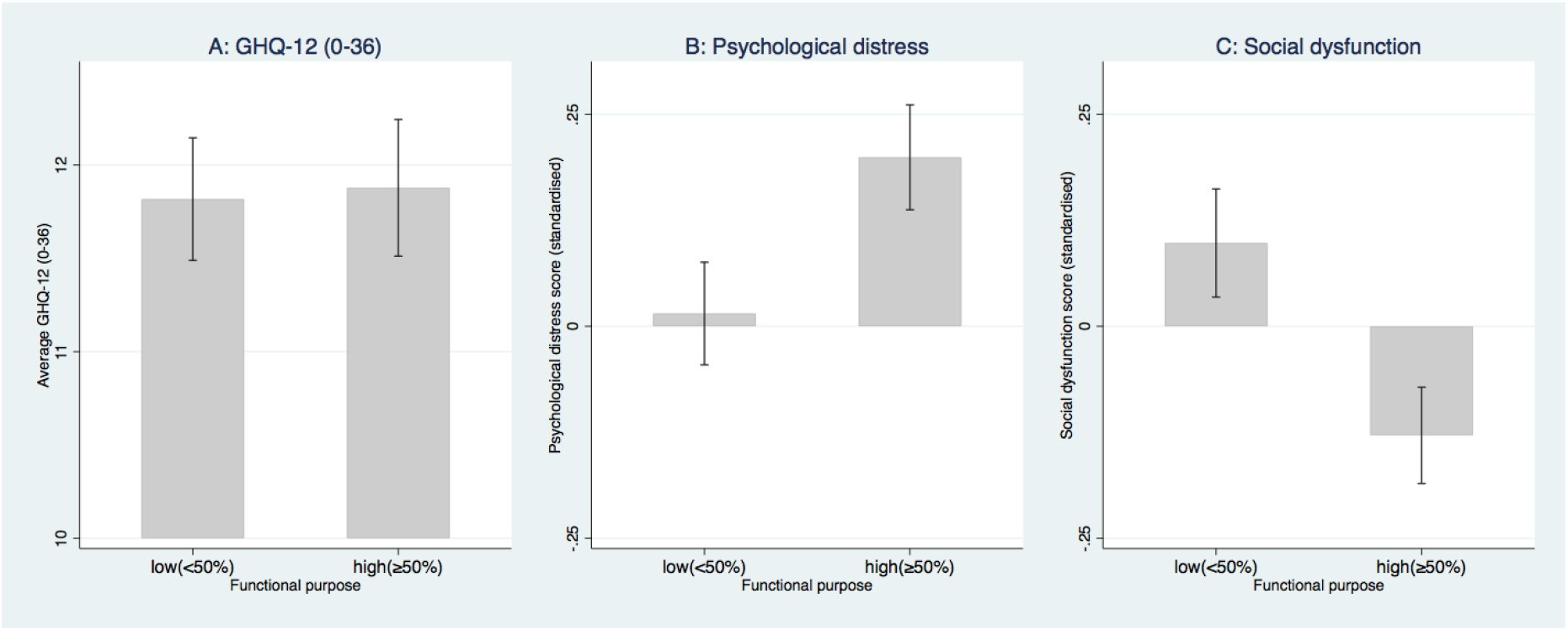
Mental health metrics by functional purpose use of the Internet

**Figure 3:**
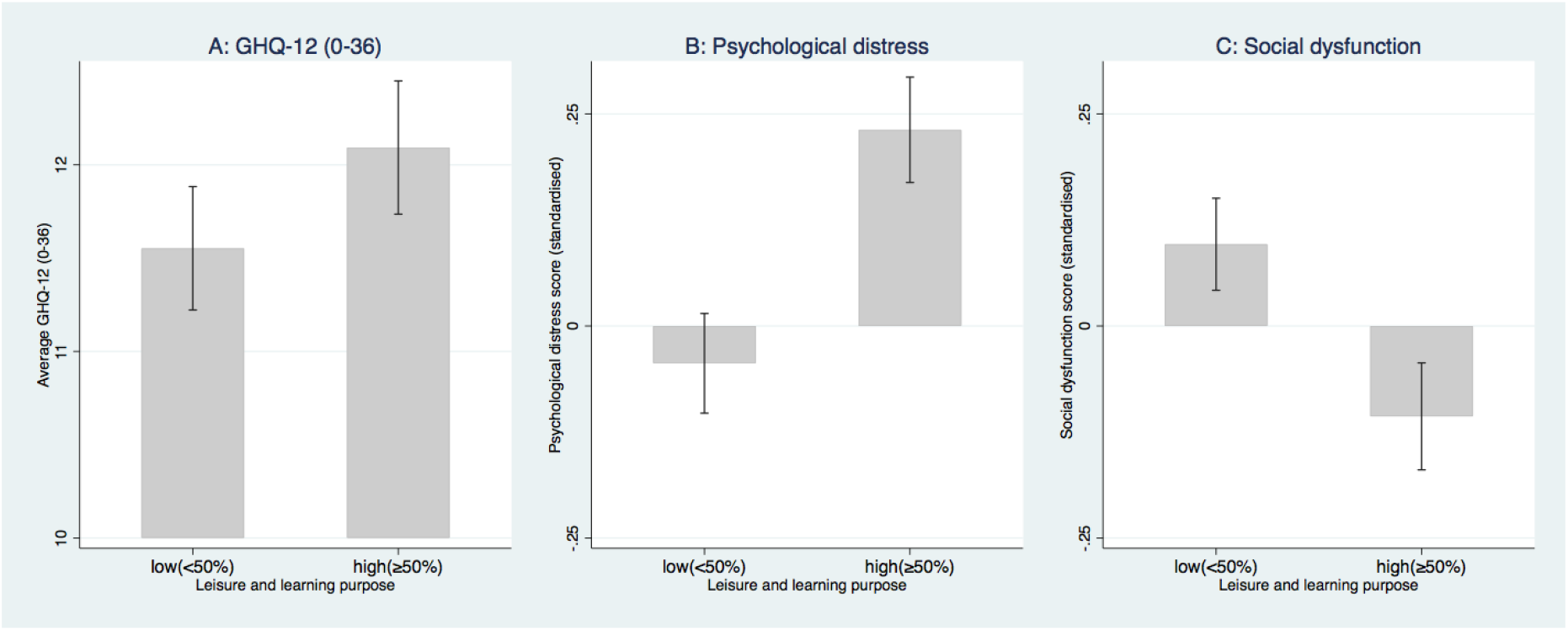
Mental health metrics by leisure-and-learning purpose use of the Internet

**Figure 4:**
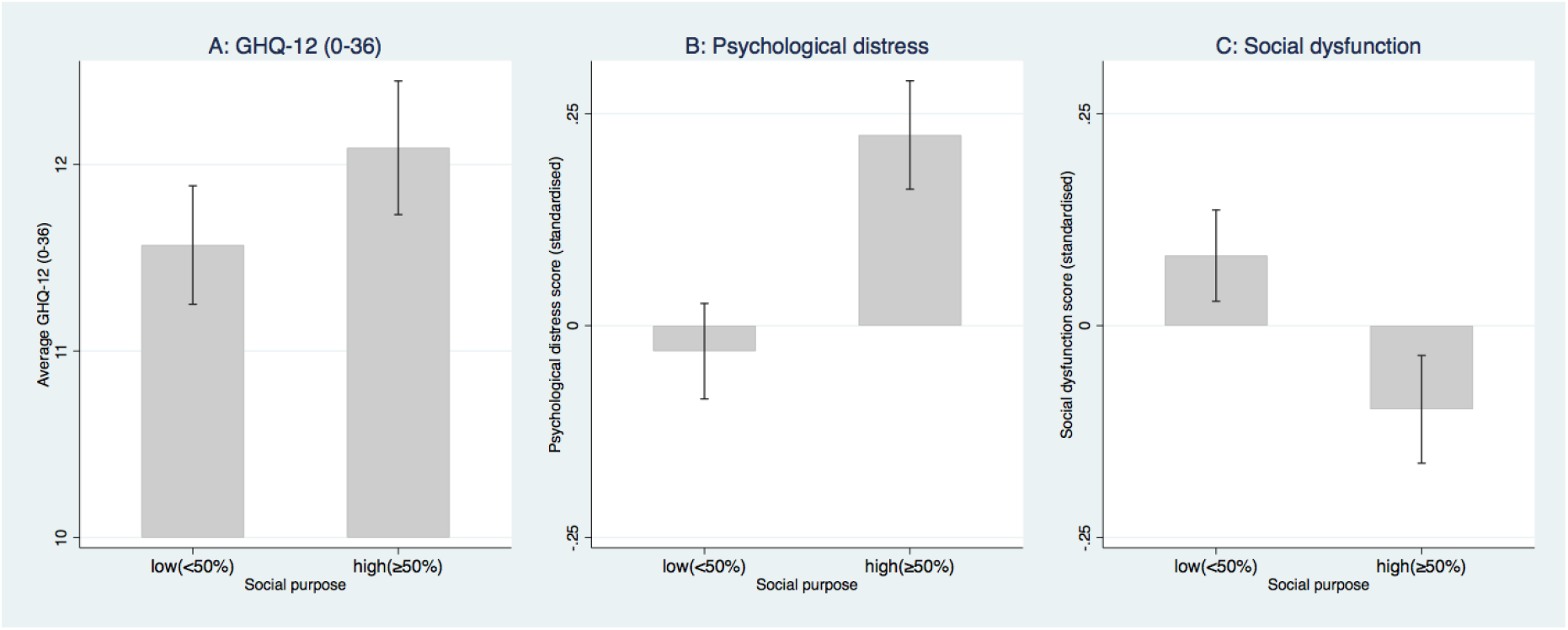
Mental health metrics by social purpose use of the Internet

## 3 Regression analysis

In this section we conduct a regression analysis to investigate: (1) whether age and gender predict internet use frequency and internet use purpose; (2) whether internet use frequency and internet use purposes predict mental health; and (3) whether such predictability varies by age and gender. We classify respondents whose age is equal or larger than the median age of the sample (59-year-old) as older respondents, otherwise we classify them as younger respondents.

### 3.1 Do age and gender predict internet use frequency and purpose?

In Table 3 we investigate whether age and gender predict internet use frequency. The table displays the regression results of a logit of internet use frequency (1 if high use –more than twice a day– vs. 0 if low use – twice a day or less). There is no significant difference between female and male in the Internet use frequency. For older individuals (≥ 59 years), the odds of high use vs. low use are 0.32 times lower than for younger individuals (*<* 59 years), given the other variables are held constant. Hence, older individuals have a lower internet use frequency, which is consistent with previous studies (Lee et al., 2011; Schehl et al., 2019).

**Table 3:** Do age and gender predict internet high use frequency? (Logit)

|  | (1)<br>Odds Ratio | (2)<br>Odds Ratio | (3)<br>Odds Ratio | (4)<br>Odds Ratio |
| --- | --- | --- | --- | --- |
| Female | 0.893<br>[0.739,1.079] |  | 0.856<br>[0.610,1.200] | 0.889<br>[0.631,1.253] |
| Age $\geq 59$ | | 0.209***<br>[0.170,0.258] | 0.218***<br>[0.158,0.301] | 0.321***<br>[0.223,0.462] |
| Female × Age $\geq 59$ | | | 0.906<br>[0.603,1.362] | 0.916<br>[0.621,1.351] |
| Controls | No | No | No | Yes |
| No. of Observations | 5980 | 5980 | 5980 | 5980 |

Note: Exponentiated coefficients are presented. Survey design and sample weights are accounted for. 95% confidence intervals in brackets. \* $p < 0.10$ , \*\* $p < 0.05$ , \*\*\* $p < 0.01$

We then investigate whether age and gender predict internet purposes by means of linear regression. Given the other variables are held constant, Table 4 Panel A shows that the functional purpose score is 0.12 standard deviations larger among younger females than among younger males, 0.42 standard deviations smaller among older males than among younger males, and 0.65 standard deviations smaller among older females than among younger males. Also, Table 4 Panel B shows that the leisure-and-learning purpose score is 0.44 standard deviations smaller among younger females than among younger males, 0.92 standard deviations smaller among older males than among younger males, and 1.18 standard deviations smaller among older females than among younger males, given the other variables are held constant. Finally, Table 4 Panel C shows that the social purpose score is 0.12 standard deviations larger among younger females than among younger males, 0.55 standard deviations smaller among older males than among younger males, and 0.41 standard deviations smaller among older females than among younger males, given the other variables are held constant.

**Table 4:** Do age and gender predict internet use purpose? (OLS)

|  | (1)<br>Coef.<br>(N=5980) | (2)<br>Coef.<br>(N=5980) | (3)<br>Coef.<br>(N=5980) | (4)<br>Coef.<br>(N=5980) |
| --- | --- | --- | --- | --- |
| **Panel A. Dependent variable: Functional purpose score** |  |  |  |  |
| Female | -0.043<br>[-0.167,0.080] |  | 0.066<br>[-0.041,0.174] | 0.121**<br>[0.010,0.231] |
| Age $\geq 59$ | | -0.908***<br>[-1.028,-0.788] | -0.718***<br>[-0.889,-0.546] | -0.415***<br>[-0.578,-0.252] |
| Female × Age $\geq 59$ | | | -0.368***<br>[-0.608,-0.127] | -0.354***<br>[-0.559,-0.149] |
| Controls | No | No | No | Yes |
| R-Squared | 0.000 | 0.152 | 0.159 | 0.252 |
| **Panel B. Dependent variable: Leisure-and-Learning purpose score** |  |  |  |  |
| Female | -0.348***<br>[-0.444,-0.252] |  | -0.480***<br>[-0.601,-0.358] | -0.444***<br>[-0.559,-0.328] |
| Age $\geq 59$ | | -1.051***<br>[-1.133,-0.970] | -1.168***<br>[-1.290,-1.046] | -0.916***<br>[-1.062,-0.771] |
| Female × Age $\geq 59$ | | | 0.180**<br>[0.030,0.329] | 0.176**<br>[0.038,0.313] |
| Controls | No | No | No | Yes |
| R-Squared | 0.025 | 0.226 | 0.262 | 0.337 |
| **Panel C. Dependent variable: Social purpose score** |  |  |  |  |
| Female | 0.189***<br>[0.103,0.274] |  | 0.141***<br>[0.045,0.237] | 0.124**<br>[0.026,0.221] |
| Age $\geq 59$ | | -0.714***<br>[-0.791,-0.636] | -0.720***<br>[-0.833,-0.607] | -0.549***<br>[-0.681,-0.417] |
| Female × Age $\geq 59$ | | | 0.027<br>[-0.130,0.183] | 0.017<br>[-0.131,0.165] |
| Controls | No | No | No | Yes |
| R-Squared | 0.009 | 0.121 | 0.126 | 0.164 |

Note: Survey design and sample weights are accounted for. 95% confidence intervals in brackets
\* p<0.10, \*\* p<0.05, \*\*\* p<0.01

Our analysis reveals that younger women use the Internet for social purposes more than men do, while younger men use the Internet for leisure-and-learning purposes more than women and older men do. Gender differences in the way Internet is used have been reported previously (Jackson et al., 2001). Females are more likely to engage in social online activities, while males use the Internet more for entertainment activities (Joiner et al., 2012; Dufour et al., 2016; Chen et al., 2017; Lemenager et al., 2021). That men and women have different motivations and preferences regarding internet use and purpose (Weiser, 2000) might be indeed a reflection of gender differences in the wider society (Joiner et al., 2012).

### 3.2 Do internet use frequency and purpose predict mental health?

In Table 5 we show the relationship between mental health metrics and internet use frequency. While we do not find that internet use frequency is a statistically significant predictor of either the GHQ-12 score or the psychological distress score, high internet frequency use is negatively associated with the social dysfunction score. In other words, high internet frequency use appears to be a protective factor for social dysfunction. Those who use the Internet more than twice a day score 0.085 standard deviations below in the social dysfunction score compared to those who use the Internet twice a day or less.

**Table 5:** Does internet use frequency predict mental health? (OLS)

|  | GHQ-12 score<br>(0-36) | Psychological<br>distress score | Social<br>dysfunction score |
| --- | --- | --- | --- |
|  | (1) Coef. | (2) Coef. | (3) Coef. |
| Internet high use | -0.190<br>[-0.521,0.141] | 0.021<br>[-0.033,0.074] | -0.085**<br>[-0.163,-0.007] |
| Age and Age-squared | YES | YES | YES |
| Female | YES | YES | YES |
| Controls | YES | YES | YES |
| No. of Observations | 5980 | 5980 | 5980 |
| R-Squared | 0.549 | 0.598 | 0.260 |

Note: This table presents the estimated impact of internet use frequency on mental health. 95% confidence intervals in brackets. Survey design and sample weights are accounted for. \* $p < 0.10$ , \*\* $p < 0.05$ , \*\*\* $p < 0.01$

In Table 6 we focus on the relationship between mental health metrics and internet use purposes. Neither functional purpose nor leisure-and-learning purpose predicts any of our mental health metrics. However, the use of internet for social purposes predicts mental health. In particular, a one standard deviation increase in the social purpose score is associated with decreases in the GHQ-12 score of 0.2 units and in the social dysfunction score of 0.05 standard deviations.

**Table 6:** Does internet use purpose predict mental health? (OLS)

|  | GHQ-12 score<br>(0-36) | Psychological<br>distress score | Social<br>dysfunction score |
| --- | --- | --- | --- |
|  | (1) Coef. | (2) Coef. | (3) Coef. |
| Functional purpose score | -0.025<br>[-0.173,0.124] | 0.009<br>[-0.017,0.035] | -0.024<br>[-0.064,0.016] |
| Leisure-and-learning purpose score | 0.076<br>[-0.125,0.278] | 0.010<br>[-0.023,0.043] | 0.009<br>[-0.038,0.056] |
| Social purpose score | -0.193**<br>[-0.349,-0.038] | -0.003<br>[-0.029,0.023] | -0.054***<br>[-0.094,-0.014] |
| Age and Age-squared | YES | YES | YES |
| Female | YES | YES | YES |
| Controls | YES | YES | YES |
| No. of Observations | 5980 | 5980 | 5980 |
| R-Squared | 0.550 | 0.598 | 0.262 |

Note: This table presents the estimated impact of internet use purposes on mental health. 95% confidence intervals in brackets. Survey design and sample weights are accounted for. \* $p < 0.10$ , \*\* $p < 0.05$ , \*\*\* $p < 0.01$

These findings contrast with Braghieri et al. (2021), who use a natural experiment, namely the staggered introduction of Facebook across US colleges, and find evidence that social media use has a negative causal effect on mental health among college students in the US. These divergent findings can be driven by several factors, including: differences in the populations under study (i.e., college students in the US vs. adults in the UK), differences in the empirical formulation and methodology of the analysis (e.g., indicator of Facebook availability at colleges in expansion group vs. social purpose score derived from PCA), and, perhaps more importantly, differences in the time period (our focus is on the COVID-19 pandemic period).

Finally, when interpreting the findings in Table 5 and Table 6 it is important to take into account that different demographic groups use the Internet at different frequencies and for different purposes, and that frequency and purpose can have different effects across different groups. This heterogeneity can mask underlying group-specific relationships. For this reason, in the next subsection we investigate whether the previous associations vary by gender and age.

### 3.3 Internet frequency use, internet use purpose and mental health by age and gender

We now investigate whether the relationships between internet use frequency, internet use purpose and mental health metrics vary by gender and age. This analysis is motivated by two sources of heterogeneity. First, as we have seen before, gender and age are important predictors of both internet use frequency and purpose. Second, it is well known that the COVID-19 pandemic has had a more detrimental effect on mental wellbeing among women and younger individuals than men and older individuals (Banks & Xu, 2020).

Figure 5 plots the coefficients on high internet use (more than twice a day) for men and women by age group (≥ 59 vs. *<* 59) after running separate regressions of mental health metrics by gender and age group. The figure reveals strong heterogeneities: internet use is a protective factor for mental health and social dysfunction among younger women, but a risk factor for mental health and psychological distress among younger men. More specifically, we find that, among younger women, those who use the Internet more than twice a day score (on average) 0.87 units below in the GHQ-12 compared to those who use the Internet twice a day or less. Moreover, they score 0.29 standard deviations below in the social dysfunction score. Among younger men, we find that high use of internet is associated with increases in the GHQ-12 score of 0.73 units and in the psychological distress score of 0.16 standard deviations.

**Figure 5:**
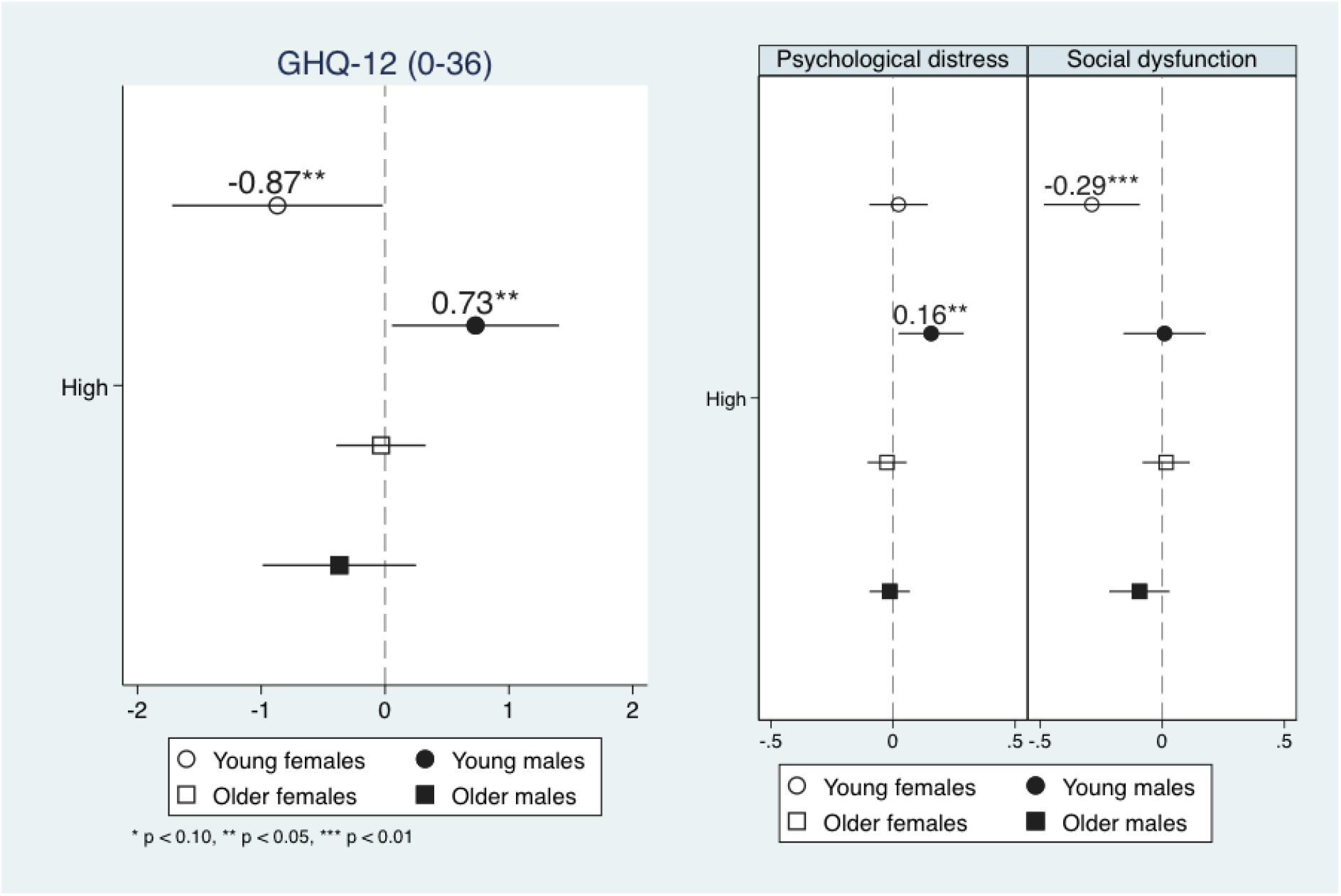
Does internet use frequency predict mental health differently by age and gender?

Figure 6 plots the coefficients on the different internet use purpose indicators (functional, leisure-and-learning, social) for men and women by age group after running separate regressions of mental health metrics by gender and age group. The figure reveals strong heterogeneities too: while leisure-and-learning purpose is a protective factor for mental health and social dysfunction among younger women, it is a risk factor for mental health and social dysfunction among younger men. More specifically, among younger women, a one standard deviation increase in the leisure-and-learning purpose score is associated with decreases in the GHQ-12 score of 0.31 units and in the social dysfunction score of 0.08 standard deviations. Among younger men, a one standard deviation increase in the leisure-and-learning purpose score is associated with increases in the GHQ-12 score of 0.50 units and in the social dysfunction score of 0.13 standard deviations.

**Figure 6:**
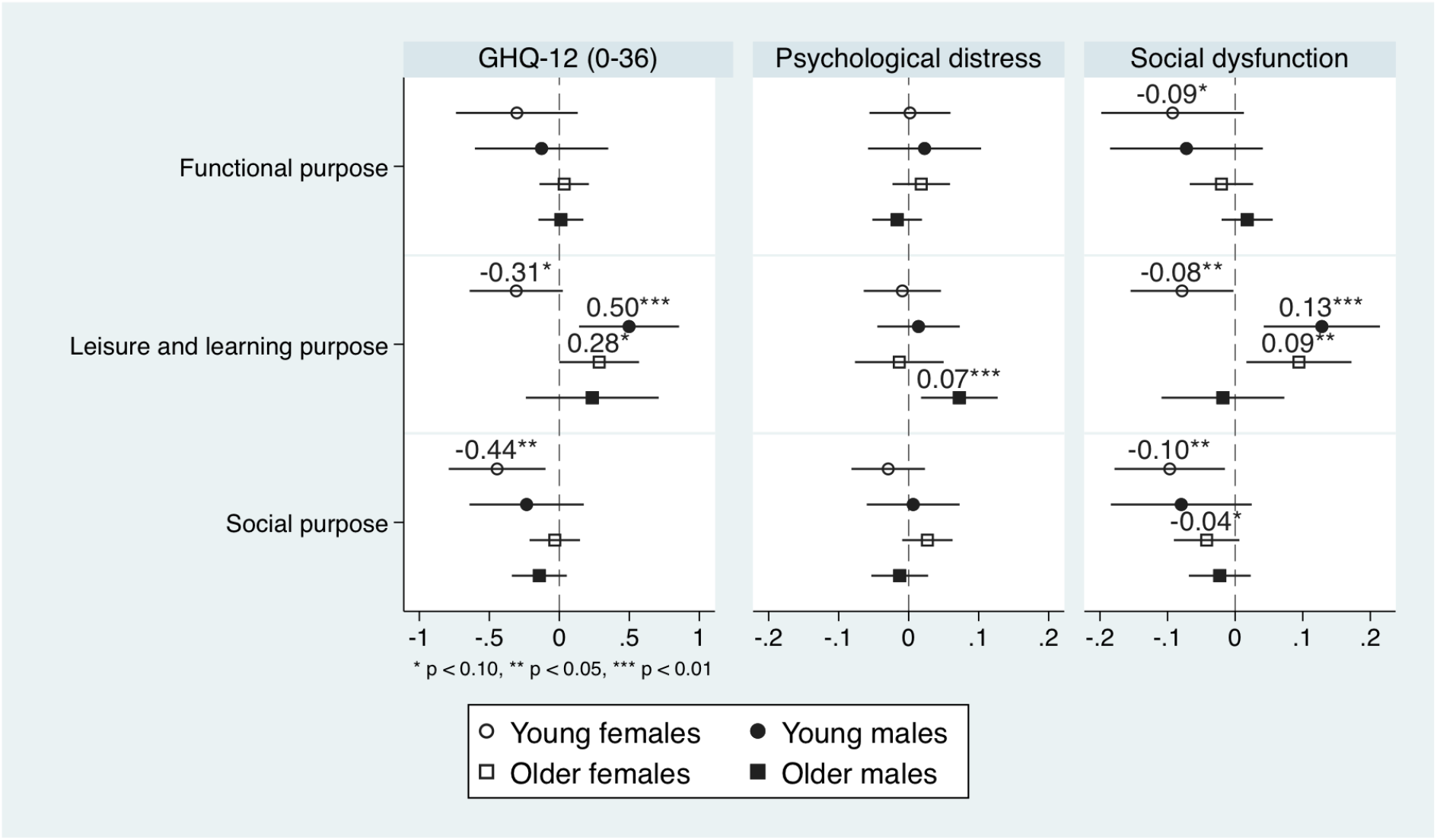
Does internet use frequency predict mental health differently by age and gender?

## 4 Statistical inference and causal interpretation

In this section we investigate whether our statistically significant findings are robust to adjusting for multiple testing, and whether our results can be given a causal interpretation above and beyond predictability.

First, we compare the p-values of individual hypothesis tests against those arising from multiple hypothesis tests. We present adjusted p-values using the step-down procedure of Romano and Wolf (Romano & Wolf, 2005, 2016) which controls for the familywise error rate (FWER). For each sample (total and group sub-samples), we use the rwolf2 command in Stata to calculate the adjusted p-values (Clarke et al., 2020; Clarke, 2021) for the coefficients on internet use frequency and purposes.

Second, and in a similar spirit to Bryan et al. (2022), who investigate the impact of mental health on the probability of being in employment for prime age workers in the UK, we implement the bounds approach proposed by Oster (2019). The key idea is to assess how important unobservable factors should be relative to observable ones in order to nullify our estimated coefficients. The estimates are denoted as *δ*, and Oster suggests a threshold of 1. We follow the suggestion by Oster to use 1.3 times the *R*^2^ value of the most extensive specification.

The results of internet use frequency on mental health, presented in Table 7, are reassuring: our previous statistically significant findings are robust to adjusting for multiple testing, and the selection on unobservable factors would have to be at least 200% as strong as selection on observables to reduce the estimated effect to zero. Finally, in Table 8, we adjust for multiple testing our p-values for the hypotheses about the relationship between internet purpose and mental health. Once again, most of our statistically significant findings are robust to adjusting for multiple testing, except for the negative association between leisure and learning purpose score and the GHQ-12 score among younger women and the negative association between social purpose score and the social dysfunction score among older women. They were significant at the 10% level and become insignificant after adjusting for multiple testing.

**Table 7:** Sensitivity tests: internet use frequency

|  | Younger |  | Older |  |
| --- | --- | --- | --- | --- |
|  | (1) female<br>(N=1756) | (2) male<br>(N=1072) | (3) female<br>(N=1693) | (4) male<br>(N=1459) |
| **Panel A. Dependent variable: GHQ-12 score (0-36)** |  |  |  |  |
| Internet high use | -0.870 | 0.730 | -0.039 | -0.379 |
| P-value | 0.045 | 0.036 | 0.862 | 0.247 |
| Adjusted p-value | 0.082 | 0.099 | 0.983 | 0.707 |
| $\delta$ | **9.62** | **2.83** | | |
| **Panel B. Dependent variable: Psychological distress score** |  |  |  |  |
| Internet high use | 0.022 | 0.156 | -0.025 | -0.014 |
| P-value | 0.719 | 0.023 | 0.561 | 0.747 |
| Adjusted p-value | 0.949 | 0.068 | 0.927 | 0.959 |
| $\delta$ | | **3.98** | | |
| **Panel C. Dependent variable: Social dysfunction score** |  |  |  |  |
| Internet high use | -0.289 | 0.009 | 0.016 | -0.094 |
| P-value | 0.004 | 0.916 | 0.753 | 0.137 |
| Adjusted p-value | 0.006 | 0.969 | 0.983 | 0.491 |
| $\delta$ | **5.00** | | | |

**Table 8:** Sensitivity tests: internet use purposes

|  | Younger |  | Older |  |
| --- | --- | --- | --- | --- |
|  | (1) female<br>(N=1756) | (2) male<br>(N=1072) | (3) female<br>(N=1693) | (4) male<br>(N=1459) |
| **Panel A. Dependent variable: GHQ-12 score (0-36)** |  |  |  |  |
| Functional purpose score | -0.304 | -0.127 | 0.035 | 0.011 |
| P-value | 0.179 | 0.613 | 0.708 | 0.895 |
| Adjusted p-value | 0.327 | 0.965 | 0.983 | 0.959 |
| Leisure-and-learning purpose | -0.308 | 0.498 | 0.285 | 0.235 |
| P-value | 0.069 | 0.008 | 0.049 | 0.337 |
| Adjusted p-value | 0.124 | **0.021** | 0.114 | 0.827 |
| Social purpose score | -0.445 | -0.234 | -0.032 | -0.143 |
| P-value | 0.010 | 0.273 | 0.733 | 0.172 |
| Adjusted p-value | **0.015** | 0.665 | 0.983 | 0.521 |
| **Panel B. Dependent variable: Psychological distress score** |  |  |  |  |
| Functional purpose score | 0.002 | 0.023 | 0.018 | -0.016 |
| P-value | 0.952 | 0.592 | 0.399 | 0.375 |
| Adjusted p-value | 0.949 | 0.965 | 0.860 | 0.827 |
| Leisure-and-learning purpose score | -0.009 | 0.014 | -0.013 | 0.072 |
| P-value | 0.751 | 0.656 | 0.667 | 0.013 |
| Adjusted p-value | 0.949 | 0.965 | 0.983 | **0.031** |
| Social purpose score | -0.029 | 0.006 | 0.027 | -0.013 |
| P-value | 0.279 | 0.853 | 0.170 | 0.546 |
| Adjusted p-value | 0.481 | 0.969 | 0.393 | 0.900 |
| **Panel C. Dependent variable: Social dysfunction score** |  |  |  |  |
| Functional purpose score | -0.093 | -0.072 | -0.020 | 0.018 |
| P-value | 0.089 | 0.214 | 0.405 | 0.370 |
| Adjusted p-value | 0.146 | 0.585 | 0.860 | 0.827 |
| Leisure-and-learning purpose score | -0.079 | 0.128 | 0.094 | -0.018 |
| P-value | 0.052 | 0.004 | 0.019 | 0.704 |
| Adjusted p-value | **0.082** | **0.013** | **0.030** | 0.959 |
| Social purpose score | -0.097 | -0.080 | -0.042 | -0.023 |
| P-value | 0.018 | 0.139 | 0.087 | 0.339 |
| Adjusted p-value | **0.036** | 0.416 | 0.229 | 0.827 |

## 5 Conclusion

This paper has documented several robust findings about the relationship between internet and mental health during the COVID-19 pandemic in the UK. Generally, high frequency internet use appears to be a protective factor for social dysfunction and the use of internet for social purpose appears to be a protective factor for social dysfunction. However, we find heterogeneous relationships across age and gender groups.

First, no significant relationship is found between high frequency internet use (more than twice a day) and mental health among older respondents (aged 59 and above). Second, among younger respondents, high frequency internet use is a protective factor for social dysfunction in women, but a risk factor for psychological distress in men. Third, among older respondents, we find that using the Internet for leisure-and-learning purposes more often is a risk factor for psychological distress in men, and a risk factor for social dysfunction in women. Fourth, while leisure-and-learning purpose is a protective factor for social dysfunction among younger women, it is a risk factor for social dysfunction among younger men. Finally, social purpose is a protective factor for social dysfunction among younger women.

Our analysis suggests that only younger women are benefiting from using the Internet more often during the pandemic, while it is a risk factor for other groups. This finding somewhat contrasts with the finding in Golin (2021) that broadband Internet leads to worse mental health for women, but not for men, in Germany, and the finding is driven by women aged 17-30. As highlighted previously, there are several differences between Golin’s study and ours, including the fact that the period analyzed by Golin (2021) does not cover the COVID-19 pandemic.

As for older groups, our findings also differ from previous studies. During the COVID-19 pandemic, Nimrod (2020) use a random sample of 407 Israeli Internet users aged 60 years and over and find that increased Internet use for leisure (games, downloading content, websites related to hobbies, writing entries in blogs, forums etc.) is significantly associated with enhanced wellbeing in April 2020, during the lockdown. However, we find that leisure-and-learning internet use purpose is a risk factor among older respondents.

The discrepancy in these findings can be driven by multiple reasons, chief among them is the fact that these are different samples, and the fact that one survey refers to April 2020 (during lockdown in Israel) while the other to September 2020 (no lockdown in the UK). Indeed, there is longitudinal evidence from Italy (between March 12, 2020 and June 7, 2020) that the online social connections can be a protective factor from psychological distress under highly restrictive isolation (strong lockdown) conditions but not under mild isolation conditions (Marinucci et al., 2022). Recent work by Altindag et al. (in press), provides causal evidence on the negative impact of lockdowns on mental health exploiting a natural experiment in Turkey (those born in December 1955 and before were under curfew, those born in January 1956 or after were exempt).

We do not find a significant relationship between social purpose and mental health among older individuals. This is consistent with face-to-face communication not being able to be replaced by online communication (Marinucci et al., 2022). Elderly people usually interact in daycare venues, community centres, and places of worship (Armitage & Nellums, 2020). Although increased internet use for leisure may enhance a sense of social engagement, reduce loneliness and compensate for the reduced leisure repertoire during the lockdown, it might increase the time being online alone when the elderly could actually spend time outdoors (in the absence of a lockdown). While online technologies could be harnessed to provide social support networks and a sense of belonging (Newman & Zainal, 2020), they cannot replace off-line activities.

As for younger groups, our findings show strong gender differences in internet use patterns, and the relationship between internet use and mental health. Consistent with previous research, we find that males are more likely to watch videos, and listen to music, whereas females are more inclined to use communication functions and social networking services (Chen et al., 2017; Lemenager et al., 2021).

Even when individuals use the Internet for the same purpose (e.g., leisure-and-learning purpose) during the pandemic, their effects might differ by gender, and indeed we observe gender differences. The gender differences documented in this study resonate with recent research highlighting that women are more likely to focus on COVID-19 issues related to family, social distancing and healthcare, while men are more likely to focus on COVID-19 issues related to sports cancellations, the global spread of the pandemic and political reactions (Thelwall & Thelwall, 2020).

Internet use is a protective factor for mental health in women, especially in the social dysfunction dimension, perhaps by allowing them to be in touch with their close friends, family and social networks. Women may be more likely to share what video they watched or what music they listened to or conduct online leisure activities together with others. It has been found that greater use of socially-supportive coping strategies was associated with a faster rate of improvements in mental health during the pandemic (Fluharty et al., 2021). Combined with our findings, using the Internet more for social activities can be a protective factor for mental health, particularly marked amongst women. Among younger men, reducing online activity and increasing offline socializing with family and friends may be associated with better mental health.

The main advantages of our study with respect to other studies on the relationship between internet and mental health during the COVID-19 pandemic are twofold: first, the use of a large representative sample; second, the rich Internet data on both use frequency and purposes, after contributing suggested content that became the Internet use module of the September 2020 COVID-19 wave study.

While our findings may suggest the importance of considering gender-targeted prevention and intervention strategies to instruct internet use and promote mental health, our study has three key limitations that must be acknowledged. First, we use observational data in a cross-sectional setting, which limits the ability to draw causal statements. Second, the data on internet use and mental health status refer to a particular point in time during the COVID-19 pandemic, September 2020, which limits the generalization of the our findings to other time periods (Quintana-Domeque & Proto, 2022). Finally, while the GHQ-12 has been extensively validated and used in several COVID-19 related studies, it has some well-known limitations, including low predictive value (Hankins, 2008b).

## Data Availability

The research data are distributed by the UK Data Service. Researchers who would
like to use Understanding Society need to register with the UK Data Service before
being allowed to apply for or download datasets. For more information visit: https://www.understandingsociety.ac.uk/documentation/access-data
The code to replicate the analysis in this paper will be publicly available from
the Harvard Dataverse repository upon publication.

## Acknowledgments

This paper uses data from Understanding Society. Understanding Society is an initiative funded by the Economic and Social Research Council and various Government Departments, with scientific leadership by the Institute for Social and Economic Research, University of Essex, and survey delivery by NatCen Social Research and Kantar Public.

Jingya Zeng acknowledges financial support from the Economic and Social Research Council (Project Reference: 2262255).

## Data and code availability

The research data are distributed by the UK Data Service. Researchers who would like to use Understanding Society need to register with the UK Data Service before being allowed to apply for or download datasets. For more information visit the <u>link</u>.

The code to replicate the analysis in this paper will be publicly available from the Harvard Dataverse repository upon publication.

## Disclaimer

The content of this paper is solely the responsibility of the authors and does not necessarily represent the official views of the institutions they are affiliated with. All errors are ours.

## A Principal Components Analysis

**Table A1:** PCA: rotated component matrix for online activities

| Activities | Factor Loadings |  |  |
| --- | --- | --- | --- |
|  | Component1 | Component2 | Component3 |
| Browsing websites | **0.486** | -0.030 | 0.115 |
| Email | **0.546** | -0.042 | -0.005 |
| Looking at Social Media | 0.132 | -0.043 | **0.595** |
| Posting on Social Media | -0.000 | -0.050 | **0.673** |
| Online banking | **0.436** | 0.064 | -0.013 |
| Online buying | **0.463** | 0.068 | 0.001 |
| Gaming | -0.187 | 0.222 | **0.416** |
| Streaming videos | -0.031 | **0.615** | -0.001 |
| Streaming music | 0.003 | **0.592** | -0.012 |
| Education | 0.099 | **0.454** | -0.096 |

Principal component analysis (PCA) is a variable reduction technique that reduces the number of variables while retaining most of the variance of the variables (Wold et al., 1987; Abdi & Williams, 2010). We use PCA to extract the most important information from online activities. After applying PCA, variables called principal components (PC) are generated. The first PC contains most of the information of the observed variables and the second PC contains most of the information of the residual variance, and so on. To simplify the interpretation of the PC, we use Promax rotation to minimize the high loadings in each component. Promax rotation also allows PCs to be correlated. Table A1 provides the loadings in each component after Promax rotation.

## B Exploratory Factor Analysis

We use the exploratory factor analysis (EFA) (see Williams et al., 2010, for more details). EFA is different from PCA. EFA hypothesizes an underlying factor structure of a set of variables and identifies the latent constructs. Before EFA, we follow the literature testing the Kaiser-Meyer-Olkin (KMO) Measure of Sampling Adequacy and Bartlett’s Test of Sphericity to assess the suitability of our data for factor analysis. The KMO index ranges from 0 to 1. An index greater than 0.5 indicates adequate sample size for the factor analysis. The null hypothesis for Bartlett test is that variables are not inter-correlated. The KMO index of our data was 0.939. Bartlett’s test of sphericity was significant (*χ*^2^ = 87149.33, df = 66, p = 0.000).

We then use EFA and the Varimax rotation method. Varimax rotation produces factor structures that are uncorrelated and simplifies the interpretation of the factors by minimizing the number of variables that have high loadings on each factor (Williams et al., 2010). We use Kaiser’s rule to determine the number of factors in the solution (Kaiser, 1960). The Kaiser’s criteria consists in using factors with eigenvalues greater than 1. The eigenvalue is a measure of the variance of the original variables that a factor explains. If an eigenvalue is less than 1, it means that the factor explains less than a single original variable, that is, the original variable is better than the generated factor.

The EFA yielded a two-factor solution, explaining 73.73% of the total variance in items (factor 1 eigenvalue = 7.67 and factor 2 eigenvalue = 1.17). We label the factors psychological distress and social dysfunction. Items 2, 5, 6, 9, 10 and 11 correspond to psychological distress and the rest items correspond to social dysfunction (see factor loadings in Table B1). The EFA result is consistent with previous literature (Werneke et al., 2000; Montazeri et al., 2003; Hankins, 2008a).

However, we need to interpret the two-factor result as the two-dimension of the GHQ-12 cautiously. Some studies suggest that the GHQ-12 measures qualitatively different constructs (Graetz, 1991; Politi et al., 1994; Hu et al., 2007). Others suggest that the two factors identified may be resulting from positive and negative wording of the questions. In that case, the two-factor GHQ-12 is a methodological artifact which results from wording effects (Hankins, 2008a; Gnambs & Staufenbiel, 2018). Unfortunately, the number of dimensions of the GHQ-12 is still subject to an ongoing debate. We use the two-factor results in this study, which also helps to take potential response bias for positively/negatively phrased items into account.

**Table B1:** Explanatory factor analysis: rotated component matrix for the GHQ-12

| Item | Factor Loadings |  |
| --- | --- | --- |
|  | Factor1 | Factor2 |
| GHQ: concentration | 0.380 | **0.727** |
| GHQ: loss of sleep | **0.765** | 0.234 |
| GHQ: playing a useful role | 0.261 | **0.787** |
| GHQ: capable of making decisions | 0.322 | **0.819** |
| GHQ: constantly under strain | **0.823** | 0.252 |
| GHQ: problem overcoming difficulties | **0.831** | 0.346 |
| GHQ: enjoy day-to-day activities | 0.279 | **0.752** |
| GHQ: ability to face problems | 0.475 | **0.747** |
| GHQ: unhappy or depressed | **0.809** | 0.413 |
| GHQ: losing confidence | **0.815** | 0.385 |
| GHQ: believe worthless | **0.777** | 0.356 |
| GHQ: general happiness | 0.459 | **0.719** |

## C List of variables and definitions

**Table C1:** Variable Definition

|  |  |
| --- | --- |
| **Mental Wellbeing and Internet Use** |  |
| GHQ-12 score (0-36) | Self-assessed mental wellbeing (0-36 scale, higher score = poorer mental wellbeing) |
| Psychological distress score (standardised) | one dimension of GHQ-12, derived from the factor analysis |
| Social dysfunction score (standardised) | another dimension of GHQ-12, derived from the factor analysis |
| Internet high use frequency | The frequency of personal internet use. 0 if in low level (once or twice a time a day or less), 1 if in high level (several times a day or almost all of the time) |
| Functional purpose (standardised) | One of the personal internet use purposes. A component derived from the principal component analysis. Highly contributing variables are browsing websites, email, online banking and online buying. |
| Leisure-and-learning purpose (standardised) | One of the personal internet use purposes. A component derived from the principal component analysis. Highly contributing variables are streaming videos, streaming music and online education. |
| Social purpose (standardised) | One of the personal internet use purposes. A component derived from the principal component analysis. Highly contributing variables are looking at social media, posting on social media and gaming. |
| **Demographics and Socio-economic Characteristics** |  |
| Age | Age of the respondent in years |
| Female | 1 if female, 0 if male |
| Ethnic Minority | 1 if Black, Asian or other ethnic minority, 0 if White British |
| Education | 1 if higher education (university +) in 2017-2019, 0 otherwise |
| Employment status | 1 if employed (if employed and/or self-employed), 0 otherwise |
| Marital status | 1 if living with a partner, 0 otherwise |
| Household size | 1 if number of individuals in household greater than two, 0 otherwise |
| Household income | Natural log of usual weekly total household gross income |
| **Physical Health** |  |
| COVID-19 symptoms | 1 if has/had symptoms that could be coronavirus, 0 otherwise |
| Health status | 1 if has long term health condition, 0 otherwise |
| Smoking | 1 if smoking cigarettes (not including e-cigarettes), 0 otherwise |
| Drinking | 1 if drinking heavily on a 'Weekly' or 'Daily or almost daily' basis, 0 otherwise |
| Physical exercise | 1 if did moderate exercise or vigorous exercise on three days in the previous week, 0 otherwise |
| **Big Five Personality Traits in 2011-2013** |  |
| Agreeableness | Levels of agreeableness (1-7 scale, higher score = higher levels) |
| Conscientiousness | Levels of conscientiousness (1-7 scale, higher score = higher levels) |
| Extraversion | Levels of extraversion (1-7 scale, higher score = higher levels) |
| Neuroticism | Levels of neuroticism (1-7 scale, higher score = higher levels) |
| Openness | Levels of openness (1-7 scale, higher score = higher levels) |
| **Geographical Variables** |  |
| Geographic location | 12 macro geographic locations (12 binary indicators) |

## D Attrition

**Table D1:** Comparison of sample sizes and average key characteristics: September 2020 Sample vs. Final Matched Sample

|  | September 2020 Sample |  | Final Matched Sample |  | Mean Diff | Diff. P-value |
| --- | --- | --- | --- | --- | --- | --- |
|  | N | Mean | N | Mean |  |  |
| GHQ-12 score (0-36) | 10267 | 12.07 | 5980 | 11.85 | -0.22 | 0.002 |
| Internet high use frequency (0-1) | 10347 | 0.75 | 5980 | 0.74 | -0.01 | 0.198 |
| Age | 10607 | 50.25 | 5980 | 54.65 | 4.4 | 0.000 |
| Female (0-1) | 10598 | 0.53 | 5980 | 0.54 | 0.01 | 0.504 |
| Ethnicity Minority (0-1) | 10552 | 0.13 | 5980 | 0.08 | -0.05 | 0.000 |
| Higher education (0-1) | 10535 | 0.42 | 5980 | 0.47 | 0.05 | 0.011 |
| Living with partner (0-1) | 10607 | 0.62 | 5980 | 0.69 | 0.07 | 0.031 |
| Employed (0-1) | 10560 | 0.59 | 5980 | 0.60 | 0.01 | 0.024 |
| log Weekly Household Income | 8604 | 5.49 | 5980 | 5.60 | 0.11 | 0.046 |
| Has/ Had COVID-19 symptoms (0-1) | 10600 | 0.16 | 5980 | 0.16 | 0 | 0.544 |
| Smoker (0-1) | 10336 | 0.11 | 5980 | 0.10 | -0.01 | 0.022 |
| Heavily Drinking (0-1) | 10317 | 0.15 | 5980 | 0.16 | 0.01 | 0.088 |
| Regular exercise (0-1) | 9647 | 0.50 | 5980 | 0.50 | 0 | 0.847 |
| Agreeableness (1-7) | 8589 | 5.55 | 5980 | 5.55 | 0 | 0.752 |
| Conscientiousness (1-7) | 8590 | 5.47 | 5980 | 5.50 | 0.03 | 0.119 |
| Extraversion (1-7) | 8590 | 4.50 | 5980 | 4.50 | 0 | 0.698 |
| Neuroticism (1-7) | 8590 | 3.62 | 5980 | 3.58 | -0.04 | 0.138 |
| Openness (1-7) | 8589 | 4.57 | 5980 | 4.59 | 0.02 | 0.371 |
| Observations | 10607 |  | 5980 |  |  |  |

## E Means and (standard deviations) of the variables used in the analysis

**Table E1:**
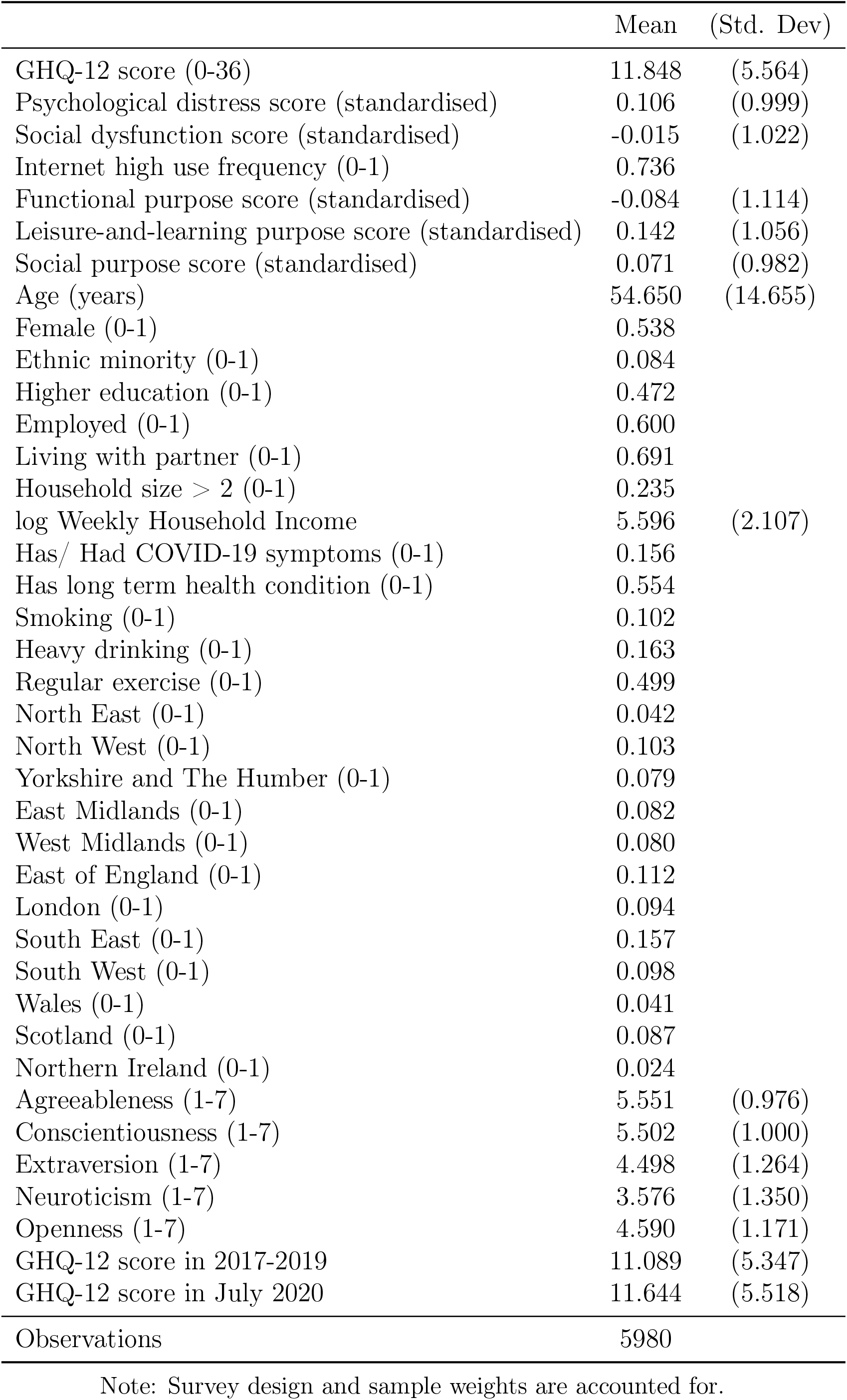
Means and (standard deviations) of the variables used in the analysis

|  | Mean | (Std. Dev) |
| --- | --- | --- |
| GHQ-12 score (0-36) | 11.848 | (5.564) |
| Psychological distress score (standardised) | 0.106 | (0.999) |
| Social dysfunction score (standardised) | -0.015 | (1.022) |
| Internet high use frequency (0-1) | 0.736 |  |
| Functional purpose score (standardised) | -0.084 | (1.114) |
| Leisure-and-learning purpose score (standardised) | 0.142 | (1.056) |
| Social purpose score (standardised) | 0.071 | (0.982) |
| Age (years) | 54.650 | (14.655) |
| Female (0-1) | 0.538 |  |
| Ethnic minority (0-1) | 0.084 |  |
| Higher education (0-1) | 0.472 |  |
| Employed (0-1) | 0.600 |  |
| Living with partner (0-1) | 0.691 |  |
| Household size > 2 (0-1) | 0.235 |  |
| log Weekly Household Income | 5.596 | (2.107) |
| Has/ Had COVID-19 symptoms (0-1) | 0.156 |  |
| Has long term health condition (0-1) | 0.554 |  |
| Smoking (0-1) | 0.102 |  |
| Heavy drinking (0-1) | 0.163 |  |
| Regular exercise (0-1) | 0.499 |  |
| North East (0-1) | 0.042 |  |
| North West (0-1) | 0.103 |  |
| Yorkshire and The Humber (0-1) | 0.079 |  |
| East Midlands (0-1) | 0.082 |  |
| West Midlands (0-1) | 0.080 |  |
| East of England (0-1) | 0.112 |  |
| London (0-1) | 0.094 |  |
| South East (0-1) | 0.157 |  |
| South West (0-1) | 0.098 |  |
| Wales (0-1) | 0.041 |  |
| Scotland (0-1) | 0.087 |  |
| Northern Ireland (0-1) | 0.024 |  |
| Agreeableness (1-7) | 5.551 | (0.976) |
| Conscientiousness (1-7) | 5.502 | (1.000) |
| Extraversion (1-7) | 4.498 | (1.264) |
| Neuroticism (1-7) | 3.576 | (1.350) |
| Openness (1-7) | 4.590 | (1.171) |
| GHQ-12 score in 2017-2019 | 11.089 | (5.347) |
| GHQ-12 score in July 2020 | 11.644 | (5.518) |
| Observations | 5980 |  |

Note: Survey design and sample weights are accounted for.

## Footnotes

1 University of Essex, Institute for Social and Economic Research, NatCen Social Research, Kantar Public (2020); University of Essex, Institute for Social and Economic Research (2020); University of Essex, Institute for Social and Economic Research (2021). Understanding Society data are available through the UK Data Service. Researchers who would like to use Understanding Society need to register with the UK Data Service before being allowed to apply for or download datasets. More information: https://www.understandingsociety.ac.uk/documentation/access-data.

2 Note that the COVID-19 survey weights map to the main wave 9 population. It provides estimates that are representative of the population of individuals (16+) resident in private households in the UK at the time of wave 9.

3 Specifically, participants in our final matched sample have (on average) better mental health (GHQ12: 11.85 vs. 12.07), are 4.4 years older on average than those in the original sample, are 5 pp less likely to be Black, Asian or from any other ethnic minority, are 5 pp more likely to have a higher education degree, are 7 pp more likely to live with a partner, are 1 pp more likely to be employed, and are 1 pp less likely to be smokers.

